# Associations between personal protective equipment and nursing staff stress during the COVID-19 pandemic

**DOI:** 10.1101/2020.08.06.20164129

**Authors:** Manuela Hoedl, Doris Eglseer, Silvia Bauer

## Abstract

**Background:** The results of several projects on the effects of personal protective equipment (PPE) have been published since the outbreak of COVID-19. It is known that wearing PPE, and specifically face masks, has physcial consequences like headache and pain, which can Increase stress among nursing staff. However, none of these studies placed a focus on PPE and nursing staff, although nurses are the only members of the health care profession who are at the patients’ bedsides 24/7, and PPE is the only way to protect them from a COVID-19 infection. Therefore, this study was carried out to investigate the association between the use of PPE and stress among nursing staff during the COVID-19 pandemic.

**Methods:** An online, cross-sectional survey was conducted, which we distributed using snowball sampling techniques. The questionnaire was developed on the basis of (inter-)national recommendations as well as the international literature. We used the perceived level of stress scale to measure the nursing staff members’ stress levels.

**Results:** We included data collected from 2600 nurses in this analysis. Nearly all nursing staff wore face masks. We showed that more than two-thirds of the nurses had moderate to high levels of stress. No statistically significant association between the use of PPE and stress was detected. However, we show a statistically significant association between the duration of mask usage and stress.

**Discussion and conclusions:** Nearly all participating nurses wore face masks or FFP masks to protect themselves from COVID-19 infection. This observation might indicate that Austrian nurses display a high level of compliance with national and international regulations and play a key role in such pandemics. Our results also show that increased mask-wearing time led to increased stress levels. These results suggest that (international regulations on how and when to use PPE should include a maximum duration of time for wearing each type of mask. Such regulations could help to prevent work-related stress, particularly in the case of future epidemics, and avoid burnout among nursing staff or even nurses leaving their jobs. The consequences of both of these negative outcomes should be considered in light of the predicted expected future shortage of health care workers.

“Contribution of the Paper”

“What is already known about the topic?”

- Associations between headache and pain experienced when wearing personal protective equipment (PPE), and specifically face masks, has already been investigated.
- Nurses are at patients’ bedsides 24/7, and PPE is the only way to protect them from a COVID-19 infection.

“What this paper adds”

- These study results show that the stress level among nursing staff during the COVID-19 pandemic ranged from moderate to high, stress levels in general, stress levels.
- We did not find a statistically significant association between the use of PPE and the nurses’ stress levels in general.
- This study identified an association between the duration of wearing PPE and the nurses’ stress levels.

## Introduction

In March 2020 the WHO assessed that COVID-19 and the underlying severe acute respiratory syndrome coronavirus 2 (SARS-CoV-2) was characterized as a pandemic. Authors of a current systematic review describe the main symptoms of COVID-19 as fever and cough followed by fatigue (1). The WHO stated on 28 June 2020 that more than 9,840,000 confirmed cases, with nearly 500,000 deaths in 216 countries/areas, had been reported (2). This pandemic has been internationally recognised as the biggest pandemic since the 1918 influenza pandemic nearly hundred years ago, during which 500 million people were infected and at least 50 million people died worldwide.

One main challenge for each affected country has been to protect high-risk groups and prevent a collapse of the health care system, and especially the intensive care system. In Austria this was done by, e.g. social distancing, working in home-office (if possible) and restricting treatments in hospitals insofar as possible.

Nevertheless, nursing staff are neither able to use social distancing nor to work in home office as preventive strategies. They have to be available at the patients’ bedsides 24/7. Therefore, it is of utmost importance to provide nursing staff with enough adequate personal protective equipment (PPE), as the virus is distributed by contact or droplet transmission. (Inter-) national organisations have launched investigations on or even created regulations for the use of PPE (3). As an example, McGilton et al. previously provided a list of considerations for infection management in the nursing home setting, which is based on (inter)national recommendations (4). They noted a need to prepare and distribute videos or other resources to nursing home staff in order to provide them with information about the adequate and correct use and disposal of PPE and to update these as needed. They also recommended that experienced nurses teach nursing home staff how to follow the PPE guidelines and how to put on and take off the PPE safely.

Several studies, letters and commentaries on PPE have been published since the COVID-19 outbreak (5-11). It is known that wearing PPE, and specifically face masks (7), can have physical consequences like headache and pain, which can increase the workload and stress levels among nursing staff over the long run. However, none of these studies placed a focus on PPE and nursing staff, although nurses are the only members of the health care profession who are at the patients’ bedsides 24/7, and PPE is the only way to protect them from a COVID-19 infection.

In addition, and specifically during the COVID-19 pandemic, an increasing number of COVID-19-affected persons can increase the stress levels among nursing staff. This is of interest because high levels of perceived stress can result in burnout and increase the risk that staff leave the nursing profession, an important consideration in light of the expected worldwide nursing shortage. Therefore, this study was carried out to describe the association between the use of different PPE and stress levels among nursing staff during the COVID-19 pandemic.

## Methods

### Design

This study employed a cross-sectional design by using an online questionnaire. The online questionnaire was distributed through the open-source, online statistical web survey app LimeSurvey. The link for the online survey was distributed by applying a snowball sampling technique with the aid of social media, such as Facebook and Twitter.

### Setting and Sample

We included Austrian nursing staff from different settings (e.g. hospital, long-term care) who worked at the bedsides of patients/residents during the COVID-19 pandemic. Some health care staff, including managers or nursing directors, were not included in the data collection process, as the aim of the study was to gain insights into bedside nursing care during this pandemic.

### Data collection instrument

Data were collected on sample characteristics such as age and gender. In addition, we collected information on the type of health care institution (e.g. hospital, long-term care, rehabilitation), professional qualifications held by the staff member (i.e. nurse, nurse aid, nursing student) and years of nursing experience (i.e. < 5 years, 5-10 years, 11-20 years, or >20 years).

The questionaire used was developed on the basis of Donabedian’s quality of healthcare model (12), which includes three levels: the structural, process and outcome levels. The questions asked on the structural and process levels were developed on the basis of official recommendations from the WHO (13,14), the Austrian Federal Ministry for Social Affairs, Health, Care and Consumer Protection (15-18), or similar guidelines extracted from international publications (7,19).

On the structural level, we asked how long the participants wore these face masks and FFP (filtering facepiece) masks, before using a new one. They could choose from among the following options: less than 4 hours, 4-8 hours, more than 8 hours, or I do not use them. On the process level, we collected data on the performance of personal protective interventions, such as the use of masks, eyewear (Yes/No). On the outcome level, we used the validated Perceived Stress Scale (PSS) to measure stress levels among the nursing staff (20). The PSS is available in the German language (21) and shows good psychometric properties. In addition, due to the fact that it only includes ten items, its use is highly practical (22). The PSS has been used internationally in several studies with different samples, including pharmacy students, informal caregivers, nursing students and nursing staff (23-32). Even though all data were collected anonymously, we obtained the required ethical approval from the responsible ethical committee.

### Data analysis

We used SPSS version 26 for data analysis (33). We expressed categorical variables as frequencies and metric variables as means. To investigate associations between the use of PPE and the staff member’s stress level, we performed a chi-square test and used the contingency coefficient (CC) as an effect size, when the tables were symmetric. In all other cases, we used Cramer’s V as a measure of the effect size. In order to validate these findings, we calculated an Kruskal-Wallis test with an independent sample for duration of wearing the mask and the PSS sum score. We considered a *p-* value of < 0.05 as statistically significant.

### Ethics

The study was approved by a responsible ethical comitee (32-386 ex 19/20). On the first page of the documentation provided, all participants were informed of the aim of the study, the responsible organisation and the contact persons as well as data security. All data collected were anonymised, and IP adresses were not stored. In addition, the data created were stored on the server of the Medical University of Graz. All participants were asked to provide their written informed consent in the first question of the online survey to comply with the General Data Protection Regulation issued by the European Union.

## Results

Five participants out of the entire sample of 2602 individuals were 65 years or older, which is above the Austrian retirement age; therefore, data from these participants were excluded from this analysis. The majority of the participating nurses worked in hospitals (73.3%), followed by long-term care institutions (17.2%). Table 1 displays the sample characteristics.

**Table 1.** Sample characteristics

|  | Nursing staff (*N* = 2597) |
| --- | --- |
| Female (%) | 83.8 |
| Mean age in years (SD) | 38 (11) |
| Professional qualification (%) |  |
| Nurse | 64.0 |
| Nurse with academic degree | 15.3 |
| Nursing aid | 11.8 |
| Nursing student | 6.5 |
| Other | 2.3 |
| Experience (%) |  |
| < 5 years | 27.1 |
| 5–10 years | 19.5 |
| 11–20 years | 20.9 |
| > 20 years | 32.5 |

Almost 80% of the participants were nurses or nurses with an academic degree. Table 2 describes the main variables of interest.

**Table 2.**
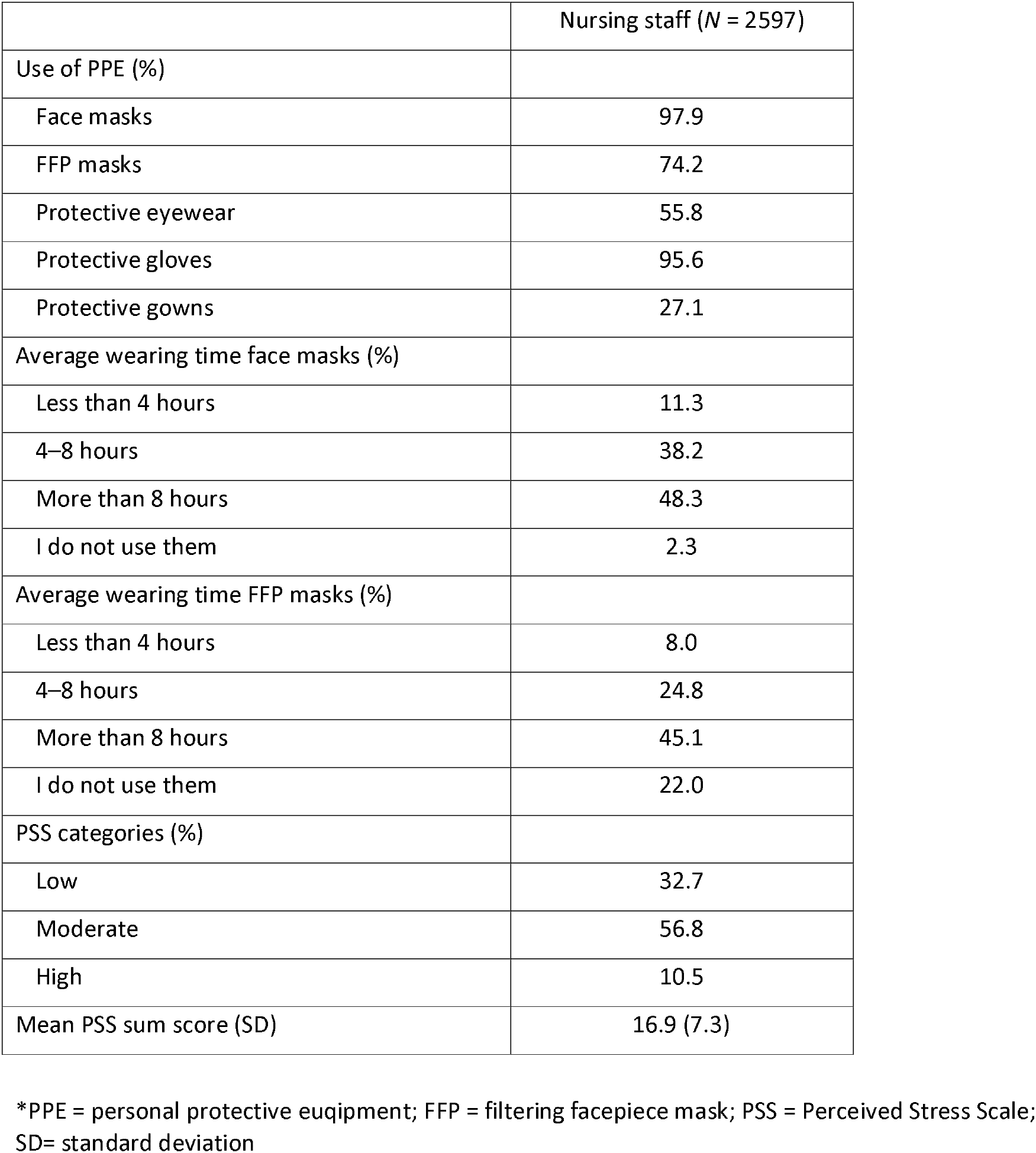
Use of PPE, mask-wearing time and perceived stress levels

Nearly all participating nurses used face masks and protective gloves during their daily work. In addition, about three-quarters of the staff used FFP masks, and 55% used protective eyewear such as glasses. Nearly half of the nursing staff wore the face masks (48.3%) and, respectively, the FFP masks (45.1%) for more than eight hours continuously. More than half of the nursing staff had moderate stress levels (56.8%), and 10.5% reported experiencing high stress levels.

Table 3 reports the association between the use of PPE and the perceived level of stress among the participating staff.

**Table 3.** Use of PPE and stress

|  | Nursing staff perceived stress level (%) |  |  |  |  |
| --- | --- | --- | --- | --- | --- |
|  | Low | Moderate | High | *p*-value of effect size | Effect size Cramer |
| Use of face masks |  |  |  |  |  |
| Yes (n= 2542) | 33.0 | 56.6 | 10.4 | 0.069 | 0.045 |
| No (n = 55) | 18.2 | 69.1 | 12.7 |  |  |
| Use of FFP masks |  |  |  |  |  |
| Yes (n = 1927) | 32.1 | 57.5 | 10.4 | 0.462 | 0.024 |
| No (n = 670) | 34.5 | 54.9 | 10.6 |  |  |
| Use of protective eyewear |  |  |  |  |  |
| Yes (n = 1436) | 32.6 | 57.2 | 10.2 | 0.831 | 0.011 |
| No (n = 1141) | 32.9 | 56.3 | 10.8 |  |  |
| Use of protective gloves |  |  |  |  |  |
| Yes (n = 2482) | 32.8 | 56.6 | 10.7 | 0.306 | 0.030 |
| No (n = 115) | 30.4 | 62.6 | 7.0 |  |  |
| Use of protective gowns |  |  |  |  |  |
| Yes (n = 1892) | 32.8 | 57.1 | 10.1 | 0.663 | 0.017 |
| No (n = 705) | 32.5 | 56.2 | 11.3 |  |  |

In general, we found no statistically significant association between the use of PPE and stress. However, one-third of the nursing staff who used face masks experienced a low stress level. In contrast, less than 20% of nursing staff who did not use face masks experienced a low stress level. Among the nurses who used gloves, 56% reported experiencing a moderate stress level. This finding differed from that for nurses who did not wear gloves, 62% of whom experienced a moderate stress level. Figure 1 shows the influence of the duration of wearing face masks or FFP masks on stress in our sample.

**Figure 4.**
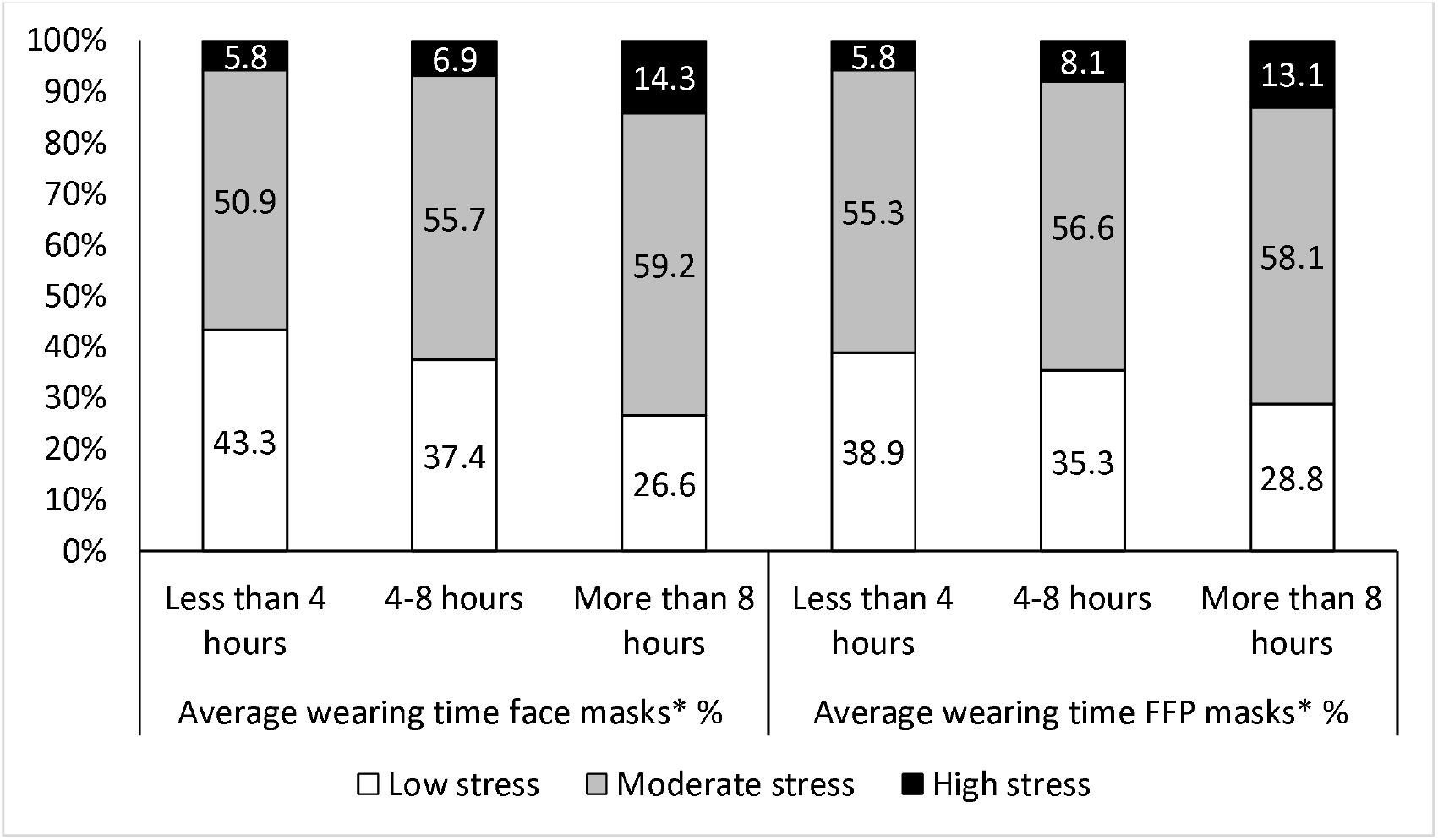
Duration of wearing face masks or FFP masks and stress (* *p*-value < 0.05)

In general, the longer the nurses wore the face masks or, respectively, the FFP masks, the higher their perceived stress level. The highest stress level was reported by nurses who wore masks for longer than eight hours (face masks 14.3%, FFP masks 13.1%). Nurses who wore face masks for more than eight hours had statistically significant higher stress levels than those who used these types of masks for less than four hours or for between four to eight hours. Nurses who wore FFP masks for more than eight hours had statistically significantly higher stress levels than those who used these types of masks for less than four hours, from four to eight hours, or did not use these masks at all.

Regardless of the types of masks (face/FFP), the perceived stress level increased statistically significantly (*p* = 0.000, CC = 0.162 vs. *p* = 0.000, CC = 0.112) as the average mask-wearing time increased. In order to validate these findings, we conducted a Kruskal-Wallis test with an independent sample for the duration of the mask-wearing time and calculated the PSS sum score. Again, we found a statistically significant difference with regard to both wearing times (face/FFP) and the perceived stress level of nurses.

## Discussion

This study was carried out to investigate the association between the use of PPE and stress among nursing staff during the COVID-19 pandemic. We could show that nearly all nursing staff wore facemasks during the study period. In addition, we show that more than two-thirds of the participants had moderate or even high-perceived stress levels. We did not identify a statistically significant association between the use of PPE and stress. However, we could show that a statistically significantly association existed between the duration of the use of masks and stress.

Nearly all staff who participated in our online survey used face masks, and nearly 75% used FFP masks. These findings contrast with those of a recent study, in which health care workers from a national university hospital in Singapore were investigated, placing a focus on PPE use and headaches (7). In this study, all participants wore FFP masks. In our study, only 55% of our participating nursing staff wore protective eyewear such as glasses. The former study placed a focus on PPE and headaches and reported that the majority of their participants wore protective eyewear (96.8%)(7).

One possible reason for the differences in the use of masks and eyewear is that most of the health care workers in the former study worked on high-risk hospital wards, such as the isolation wards, emergency rooms - including a fever facility - and the medical intensive care unit (7). In addition, the use of PPE was mandatory for all health care workers on these wards (7), which might explain why the numbers of staff wearing masks and eyewear were higher than those seen in our study. Another explanation for these differences can be that our online survey was initiated on 12 May 2020. This date was approximately two months after the lockdown was initiated in Austria and after the first official COVID-19 regulations were issued by the Austrian government. In contrast, the study from Singapore was conducted shortly after the first wave of COVID-19 cases was reported there.

Most of our study participants wore surigcal face masks and the FFP masks for more than four hours (86.5% vs. 69.9%). This finding is in line with those for the health care staff working in high-risk wards in the former study, who stated that they used the N95 face masks on average 5.9 hours each day (7). These results support the recommendations made by the National Institute for Occupational Safety and Health regarding the extended use of N95 masks (34). These recommendations state that the masks should be used for extended periods, as touching the masks less frequently might result in a lower risk of contact transmission as opposed to reusing the mask (34).

Two studies have been carried out to investigate the wearing time of N95 masks among health care workers (35, 36). In the first study, the sample included 27 health care workers, 22 of which were nursing staff, and the median wearing time for eight different N95 masks ranged between 4.1 and 7.7 hours (35), findings that are in line with our results. The second study was carried out to investigate the levels of compliance regarding mask usage and compare this with physiological effects and subjective symptoms (36). In this study, ten nurses participated, who had an average of eleven years of experience with wearing an N95 mask (36). The authors reported that the daily average wearing time ranged between 159.1 and 223.7 minutes and that 90% of the nurses tolerated the use of the mask for two 12-hour shifts (36). Even though the nurses had significant amounts of experience with wearing masks in this study, the low average mask-wearing time reported was surprising, as compared to the average mask-wearing time found in our study. This finding could be explained by the fact that our study was conducted during the COVID-19 pandemic, when the risks and consequences of wearing or not wearing the masks were onmipresent. The former study by Rebmann et al., in contrast, was performed in 2013 at a time when no worldwide pandemic was ongoing; therefore, the results are not completely comparable.

We could show that nurses who wore a face mask less than four hours and up to eight hours per day reported lower stress levels more often than nurses who wore the FFP mask for the same time span. This result agrees with the results of Rebmann et al., which show that the daily average wearing time of an N95 mask alone was higher than the combination of wearing an N95 with a mask overlay (36). This finding might be explained by the fact that a face mask in our study (the N95 mask described by Rebmann et al.) is thinner than an FFP mask in our study. In addition, these authors showed that wearing an N95 masks with an extra overlay statistically significantly increases CO_2_, nausea and visual challenges as compared to wearing a N95 mask alone (36).

Among the nursing staff, between 5.8% and 14.3% experienced high stress levels. Regardless of the type of masks (face or FFP), the perceived level of stress was statistically significant and positively correlated with the increased mask-wearing time. Other studies have generally showed - and specifically in the case of PPE usage - that mask-wearing for more than four hours per day is associated with headaches (7), increased levels of CO_2_, perceived exertion, shortness of breath, reported headaches, dizziness and communication difficulties (36). All of these effects can lead to discomfort, pain and consequently, an increase in the stress levels of nursing staff.

However, study authors have also reported that 22.1% of staff typically remove the masks due to reported discomfort, such as breathing difficulties (36). In another study, 59% of the participating nurses refused to wear the masks for more than eight hours. Their stated reasons included an intolerance to heat, pressure, or pain, dizziness, difficulties concentrating and interference with communication (35). Nevertheless, we conducted this study to provide an insight into the use of PPE in Austrian hospitals during the COVID-19 pandemic. We did not specifically assess the reasons for compliance or non-compliance with the rules for PPE use or the reasons for limiting this use, which highlights the need to carry out qualitative studies on this topic in the future.

### Strengths and limitations

The first strength of this study is that it is the first one, to our knowledge, that describes the influence of wearing personal protective equipment during the COVID-19 pandemic in Austria. Another strength is that our sample included more than 2500 nurses. One limitation of our study might be that we started the survey in mid-May, nearly two months after the COVID-19 pandemic began in Austria. Because we asked nurses retrospectively about their use of PPE during the COVID-19 pandemic, some perceptions could have been distorted.

### Conclusions

This study was carried out to investigate the association between PPE and stress levels among nursing staff during the COVID-19 pandemic in Austria. Nearly all participating nurses used face masks or FFP masks. This might be an indication of a high level of compliance among Austrian nurses regarding the national as well as international regulations and highlights the key role played by nurses in such pandemics. Our results also show that increased mask-wearing time led to increased levels of stress. These results suggest that (inter-) national regulations on how and when to use PPE should also include a maximum duration of time for wearing each type of mask. Such regulations could help to prevent work-related stress, particularly in the case of future epidemics, and avoid burnout among nursing staff or even nurses leaving their jobs. The consequences of both of these negative outcomes should be considered in light of the predicted expected future shortage of health care workers.

## Data Availability

The data can not be made available due to legal issues.

